# What do patients think about home-based testing for better asthma diagnosis? - Insights from a qualitative study

**DOI:** 10.1101/2025.08.13.25333591

**Authors:** Binish Khatoon, Joanna Smith, Stephen J Fowler, Angela Simpson, Clare S Murray, Ran Wang

**Affiliations:** Division of Immunology, Immunity to Infection & Respiratory Medicine, School of Biological Sciences, Faculty of Biology, Medicine and Health, University of Manchester, United Kingdom; School for Health and Social Care, College of Health & Wellbeing and Life Sciences, Sheffield Hallam University / Sheffield Children’s NHS Foundation Trust, Sheffield; NIHR Manchester Biomedical Research Centre, Manchester University NHS Foundation Trust, United Kingdom

**Keywords:** Asthma, home testing, variability, spirometry, FeNO, digital health

## Abstract

**Background:** Asthma is misdiagnosed in one-third of patients. Due to its variable nature, international guidelines recommend performing key diagnostic tests during symptomatic periods or in the morning to improve accuracy. Limited access to timely clinic appointments and community-based diagnostics makes this difficult. Handheld spirometry and fractional exhaled nitric oxide (FeNO) are feasible for home use, enabling timely and flexible testing.

**Objective:** To explored patients’ views on performing spirometry and FeNO at home during the asthma diagnostic process.

**Design:** A qualitative study using semi-structured interviews. Data were analysed using framework approach.

**Setting:** This prospective observational study was conducted at a National Institute for Health and Care Research Clinical Research Facility (NIHR CRF), based within a large National Health Service (NHS) Trust, as part of the Rapid-Access Diagnostics for Asthma (RADicA) study (ISRCTN11676160).

**Participants:** A purposive sample of 15 symptomatic adult patients with GP-suspected asthma who were referred for diagnostic evaluation of the condition; all patients used home spirometry and FeNO devices during their diagnostic processes.

**Results:** Three themes emerged from the analysis: ‘Perceived value of, and burdens of home testing’, ‘Views on device usability and acceptability’, and ‘Information and support needs’. Home testing was generally welcomed by patients as a way of improving their understanding of their condition and enabling an accurate diagnosis of their symptoms. Key barriers (e.g. testing frequency, lack of privacy) and enablers to improve feasibility (e.g. training and support) were also identified.

**Conclusion:** This study provides valuable insights into the barriers and enablers of home-based diagnostic strategies for asthma. Findings can inform service design and implementation approaches to enhance the feasibility and effectiveness of home testing.

**Funding:** This study was funded by National Institute for Health Research (NIHR) Research for Patient Benefit Grant (NIHR203591) and supported by the Manchester NIHR Biomedical Research Centre (BRC) (grant no. BRC-1215-20007, and NIHR203308), Asthma UK/Innovate (grant no. AUK-PG-2018-406) and North West Lung Centre Charity. The views expressed are those of the author(s) and not necessarily those of the NIHR or the Department of Health and Social Care.

**Strengths and limitations of this study:**

- A key strength to this study is that the patients participated in the interviews were representative of those at the diagnostic stage.
- Patients who participated in the interviews openly share their experiences in the absence of the main clinical research staff.
- The identified enablers and barriers to ambulatory testing using handheld devices for asthma diagnosis are transferable to other settings, including for disease monitoring and self-management.
- The study findings may not be representative of the experiences and views of all patients with suspected asthma due to the qualitative nature of the study design, but nonetheless, the current study included a diverse group of patients with broad baseline characteristics, providing useful insights into their views.

## Introduction

Over 160,000 individuals in the UK are diagnosed with asthma each year ^1–3^, but 1 in 3 are misdiagnosed. Misdiagnosis in asthma contributes to avoidable morbidity, mortality and burden on the healthcare resources. ^4–7^

Asthma is characterised by recurring symptoms of wheeze, breathlessness, chest tightness and cough caused by variable airflow obstruction, typically associated with airway inflammation. ^8^ Asthma symptoms, lung function and airway inflammation show diurnal variation and often worsen overnight or early in the morning. ^9–12^ Asthma symptoms also vary from day to day and on exposure to triggers e.g. exercise, weather change or exposure to sensitised aeroallergens. ^10^ Despite this, clinical practice currently relies on a ‘one-off’ testing approach, ^8, 11^ with tests performed at a pre-scheduled clinic regardless of whether patients are experiencing their typical symptoms at the time. It is noteworthy that spirometry and fractional exhaled nitric oxide (FeNO) measurements also demonstrate marked temporal variation even during clinical hours (typically between 9am and 5pm), with higher likelihood of a positive bronchodilator reversibility (BDR) and FeNO if tests are performed in the morning. ^9, 13^ Therefore, perhaps it is not surprising that neither single tests nor combinations of tests, performed during a planned clinic in primary care settings, were sufficiently effective in ruling in or ruling out asthma ^8, 11 14–16^. Indeed, the National Institute for Health and Care Excellence (NICE) estimates that over 60% of individuals with suspected asthma will require a referral for bronchial challenge testing^8^ - a test available only in some secondary care settings. To enhance diagnostic performances of GP-accessible tests (such as spirometry, BDR and FeNO), the Global Initiative for Asthma guidance ^11^ recommends performing these tests during symptomatic periods or in the morning^9, 10, 12^. Nevertheless, implementing this is challenging in routine care due to limited access to timely appointments.

Therefore, improving asthma diagnosis requires innovative approaches that account for test timing. ^10, 15^ Peak expiratory flow (PEF) measurements have, until recently, been the only domiciliary test available. ^8^ However, PEF is far less sensitive than forced expiratory volume within 1 second in detecting small airway obstruction; ^17^ recent evidence suggests that PEF diurnal variability has a sensitivity of only 15% for asthma diagnosis. ^18^

The UK’s 10-Year Health Plan^19^ prioritises digital healthcare and a shift from hospital to community-based care, and acknowledge respiratory health as a key focus for the coming years. The introduction of digital handheld spirometry and FeNO devices for home-based testing aligns with the drive to capitalise on the digital revolution. While both home spirometry and FeNO have shown feasibility in asthma diagnosis and self-management, ^20–22^ understanding their acceptability for diagnostic use is a critical step toward real-world implementation.

In this study, we explored patients’ views on the potential benefits, barriers and enablers of home-based testing using handheld spirometry and FeNO during the asthma diagnostic process.

## Methods

### Study and design

A qualitative descriptive design was adopted, as qualitative methods were appropriate to explore and describe patient’s perspectives and experiences of home-based testing for asthma diagnosis. All participants received a written information sheet and gave written informed consent before the interview. The study methods and results are reported according to the Consolidated Criteria for Reporting Qualitative Research guidelines This study was approved by the Research Ethics Committee (18/NW/0777; ISRCTN 11676160).

### Participant recruitment and procedures

The study is nested in the Rapid Asthma Diagnostic Clinics for Asthma study (RADicA, https://www.radica.org.uk). ^23^ Symptomatic and untreated adults (>16 years) with GP-suspected asthma were recruited from primary care, and provided with home spirometry (MIR Spirobank Smart, Intermedical, UK) and FeNO (NOBreath, Bedfont, UK) devices. With permission, the mobile software application (‘app’) associated with the spirometers was downloaded, set up and installed on the participants’ smartphones in the RADicA clinic during their appointments. The app software provides a virtual assistant for users to achieve optimum spirometry blow technique. In-person training and written instructions were provided for both devices; short explanations about what FeNO and spirometry measure and simple guides were given to participants regarding what the results may mean.

Participants were asked to take the devices home and take 2-4 measurements a day for two weeks (first stage of the study, see online supplement). A total of 51 patients participated in the first stage of the study that assessed the adherence and feasibility of ambulatory asthma testing.

For this qualitative stage, participants from the study cohort were invited to take part in a semi-structured interview with BK, an experienced qualitative researcher, at the end of stage 1. Participants were approached and participant information sheet given in person during face-to-face device training prior to the monitoring period. Invited participants were then re-contacted by BK via telephone or in person following monitoring period for consenting into the interviews. With participants’ permission the interviews were audio recorded.

### Data collection

The interviews were undertaken between Aug 2023 and May 2024 at the RADicA study clinic office in Manchester University NHS Foundation Trust, Greater Manchester, UK. The 1:1 interviews were undertaken in person or by telephone depending on participants’ preference and lasted between 30-45 minutes; all interviews were conducted in the absence of the RADicA clinical research team, allowing open sharing of their experiences and views.

A topic schedule, developed prior to the commencement of the study (online supplementary file) and informed by patient and public involvement workshops and feedback, the study aims and review of literature, was used to guide the interview. BK, who is independent of the RADicA clinical team, conducted all interviews, building rapport with participants during the consent process, and at the start of each interview when this stage of research was further explained.

Study recruitment finished when BK, in discussion with JS, recognising participants were not offering any additional information and the data are of sufficient depth to meet the study aims.

### Data analysis

The framework approach was used to analyse the interview data^24, 25^, consisting of the following stages:

1. Data management: recorded interview data and field notes were transcribed verbatim by BK and uploaded into the qualitative data software package NVivo12. BK undertook the initial coding, assigning ‘labels’ to data extracts, before bringing these together into preliminary sub-themes, with discussions with JS, the qualitative research adviser (independent to the clinical research team), throughout.
2. Descriptive accounts: once all the transcribed data were assigned into preliminary sub-themes, the initial subthemes were refined and associations between sub-themes were identified. At key stages, during the iterative development of sub-themes, they were discussed with the wider research team, an essential part of refining and developing broader final themes. If there were any disagreements during this phase, team meetings were held to discuss and to come to an agreement.
3. Exploratory accounts: final themes and sub-themes were agreed at team meetings, resulting in a cohesive narrative that met the overall study aims, and ensured that the findings reflected the perceived benefits, burdens and potential barriers of using home-based spirometry and FeNO for asthma diagnosis.

### Patient and public involvement

A Patient Advisory Group (PAG) comprising five members with asthma or caring for someone with asthma. Members of PAG contributed to the inclusive research recruitment strategy, the lay summary of the research proposal and provided feedback on study design. They also reviewed and provided feedback on all patient-facing materials, including interview topic guides (Table E1), symptom and test diaries, device training materials and instructions, participant information sheets, and consent forms. One PAG member also served on the trial steering committee.

## Results

Of 26 invited individuals, 16 patients were interviewed with one participant’s recording corrupted and therefore not usable; two did not attend the appointment and 8 declined without giving a reason. Data collected from 15 participants were included in the analysis. The three overarching themes that emerged from the analysis were, ‘Perceived values and burdens of home asthma testing’, ‘Views on device usability and acceptability’, and ‘Information and support needs’ (Table 2).

**Table 1:** Participant characteristics.

| Participant characteristics | (n = 15) |
| --- | --- |
| *Gender male: female* | 7:8 |
| *Diagnoses:* |  |
| Asthma (n) | 11 |
| Not asthma (n) | 4 |
| *Age (years)* | Range: 21-52 |
| 18-30 | 5 |
| 31-40 | 5 |
| 41-50 | 1 |
| Above 50 | 4 |
| *Ethnicity* |  |
| White British | 10 |
| Black Caribbean | 1 |
| Asian | 1 |
| Pakistani | 1 |
| Chinese | 1 |
| Africa | 1 |
| *Employment / role* |  |
| Student | 3 |
| Retired | 3 |
| Management / executive | 2 |
| Professional (teacher) | 1 |
| Sales | 1 |
| App developer | 1 |
| Security | 1 |
| Manual worker (gardener) | 1 |
| Unemployed | 1 |
| Carer | 1 |

**Table 2.** Themes and subthemes.

| Themes | Sub-themes |
| --- | --- |
| 1. Perceived values of, and burdens of home asthma testing | Motivators to perform home testing |
|  | Opportunity for condition self-management |
|  | Limitations of home testing |
| 2. Views on device usability and acceptability | Acceptability and usability of FeNO device |
|  | Acceptability and usability of spirometry device and app |
|  | Suggestions for improvement |
| 3. Information and support needs | Information provision and modes of delivery |
|  | Building confidence and on-going support |

### THEME 1: Perceived values of, and burdens of home asthma testing

A narrative throughout participants’ accounts was their *motivation for undertaking home testing,* and the opportunity for *self-managing their condition*, along with descriptions of *the limitations of home testing*.

*Motivators to perform home testing:* a desire to better understand asthma and asthma-related symptoms was a common motivator to undertaking home testing. Some participants expanded further explaining that improved understanding of their condition and the meaning of test results would help them to know how well their lungs were functioning and therefore help them manage their condition. For most participants there was recognition that home testing could contribute to the accurate diagnosing of asthma, and participating in the study could help future patients. Examples of participants’ motivators included:

> ‘I feel like healthcare has an emphasis on dealing with problems once they’ve happened, whereas for me I think I’m very keen to look at sort of, you know, these little diagnostic tests that one can do before you get ill to find out if things are worsening or if things are beginning etc. It’s a way of learning more by doing the tests so you can categorically say for people in future that they can, they’re asthmatic or not basically’ *participant 7*

> ‘I think the most important thing was whether or not this would help the doctors give me a more accurate diagnosis from my home monitoring,’ *participant 14*

*Opportunity for condition self-management* while linked to the above sub-theme more explicitly related to participants’ desires to be more proactive in self-managing their condition. They perceived this could be facilitated by home testing, through sharing and discussing results with health professionals. Some participants highlighted that home testing would empower them to act on their symptoms. Examples include:

> ‘Understanding the condition and then seeing, well, does anything need to be done? Or do I need to do something differently about the management of the condition?’ *participant 2*

*Limitations of home testing*: while sharing results could help to self-manage their condition, for some this responsibility was overwhelming, and they perceived some tasks unnecessary or overly complicated. For example:

> ‘It’s not just the instructions, I think it’s more of like when you’re doing it and passing the information over through the internet. … And I don’t think you’d be getting the right readings because you’re doing something else rather than just concentrating on doing that, you know what I mean? Then sending it off. But if somebody was, say either on the phone or you’re doing it like through now and again a home visit or something, but, which contradicts the situation of doing it from home and….’ *Participant 9*

Similarly, some participants’ perceived that whilst asthma was a long-term condition it was non-life threatening, and that symptoms were to be tolerated. Therefore, they questioned the value of home testing if symptoms were unlikely to improve:

> ‘Because this thing is when people have asthma their condition is not going to improve like dramatically overnight. Like it’s a long-term thing. So they kind of get numb with their condition. They don’t think its anything so life threatening. That’s why this kind of task which takes time, takes effort becomes a burden not something they see benefit anymore. So if its more life-threatening thing it’s definitely very, very important, like heart disease you know heart attack or cancer. With asthma it’s kind of minor people don’t really put such weight on it…,’ *Participant 4*

For other participants a drawback of home testing was an increased visibility of having asthma/ asthma symptoms, with a perception this could be stigmatising potentially impact on their mental well-being. As a consequence this may influence undertaking the tests outside the home if they could not be done in private, for example:

> ‘It’s the mental weight that comes with it. There’s kind of a stigma when for instance, so if you’re in the home, other people have probably got to be there whilst you’re doing your test and whatnot, which-It makes you feel like you’re being-Sticking out like a sore thumb. And if you happen to be out and about and you have to do it in public for instance, …. it’s going to be detrimental to them in the long run as well, so that’s the kind of element I’d look at. Other people’s opinions and whatnot. And then obviously they might not want other people to know what’s going on as well, so it’s a privacy thing as well.’ *Participant 5*

### THEME 2: Views of device usability and acceptability

Participants shared their views of the *usability and acceptability* of both devices and the associated spirometer app, *along with suggestions for improvement*. For many participants both devices were, once they became familiar with them, relatively easy to use, for example:

> ‘I found it easy. Once you’d done the initial few, many of them and obviously, it’s straight forward.’ *Participant 3*

However, there were differences in the usability of the devices as outlined below.

*Acceptability and usability of the FeNO device:* participants reported that the FeNO device was easier to use compared to the spirometry device, which typically related to the breathing technique required to undertake the test and obtaining a result with a single effort. For example:

> ‘I found that (FeNO) far easier and straightforward. But obviously, you’ve still got to master the technique, and maybe the technique is easier too, because you’ve got to get that flow just right.’ *Participant 6*

*Acceptability and usability of spirometer device and app:* portability was the main benefit of the spirometer device. A frequently reported limitation of the spirometer device was the number of times participants had to use the device to get an accurate reading; concerns about obtaining incorrect readings caused anxiety and frustration for some participants. For some participants completing the tests accurately appeared burdensome, with reports of feeling tired and frustrated if there had been many attempts to complete to get an accurate reading. Examples of the challenges reported include:

> ‘Because of more and more unsuccessful attempts …I started to wonder is there something wrong with the device and I just get some help and they tried to clean it and maybe there’s saliva wet inside. Just feel very frustrated because I need to get it done I feel like it’s a responsibility and also like a task but I can’t get a successful attempt and I have to repeat and my mouth’s very dry.’ *Participant 4*

> ‘I was just gasping for air. I’d blow into it, and I’d just be trying to catch my breath back because it just took a lot of energy out of me blowing into it, because you have to obviously hold your breath, and then do a long breath, …. I found it very difficult.’ *Participant 1*

The spirometer device was linked to an app, which in general participants reported the functions were easy to use:

> ‘The app was great, so easy to use. They showed me how to use it anyway. And it was just, that was all right. It’s dead easy to use. But like I’ve got an iPhone so it’s just quite simple to just send the results over.’ Participant 11

While some participants highlighted the app was self-explanatory and the instructions were simple and easy to follow, others struggled to remember how to use the app and had technical difficulties. For example:

> ‘I found that a bit more difficult to use in terms of it was a lot more complex, like the process. Because obviously, you had to get that app up. And also, I found it hard….I was doing my results a week and a half into it, they wouldn’t accept any of the results I tried to submit….I was paranoid that the team would have thought that I wasn’t doing it properly, I wasn’t doing what I was meant to do. But they were aware that the app was down, so it wasn’t that bad.’ *Participant 1*

*Suggestions for improvement*: Participants suggested FeNO devices could be redesigned for home use and uploading results via the app could be instantaneous to health professionals rather than downloading and attaching to an email. In addition, icons on the spirometry app could be simplified, including a clear explanation of their meaning. For example:

> ‘Surely there’s a way in the app where you can have it link up like so it’s almost like on a cloud service where you guys can see the results straight away rather than having to send an attachment. That was absolutely fine but like I’m sure, I’m sure there are ways you can have something on a cloud of some sort like just online and it’s like updated. That would just you know save a bit of time doing it because that was twice a day wasn’t it that one?’ *Participant 13*

> ‘…The pictures on the screen, I think it looks like it’s very small. I think the biggest picture was about yea big, which-If anybody’s hard of sight, regardless of age again, not going to be the most friendly and especially if they’ve got any memory issues as well….’ *Participant 5*

While overall participants reported the FeNO device to be simple and easier to use compared to the spirometry device, participants reported the size of the device and having to change the tubes made is difficult for use outside the home, for example:

> ‘The purple one (FeNO) was absolutely fine, I mean the only thing that was a bit of a hassle with swapping out the thing, remembering to swap out the tubes … it was just big’ *Participant 13*

### THEME 3: Information and support needs

Participants highlighted that information and support were vital to the success of home testing, which included the way *information is provided* and *building confidence* and *on-going support*.

*Information provision and modes of delivery:* participants perceived that the training sessions were supportive, unrushed, carried out in a friendly environment and information provided was sufficient to prepare them to complete the tests at home independently, for example:

> ‘Actually, at the hospital, when they handed over both devices, they showed me how to do it. I came back. I started doing it like, you know-It was easy and even, you know, in the book, the manual, at the hospital, I didn’t even check the manual because, you know, it was very easy.… the lady at the hospital, she showed me how to do it, how to use it…I was like, “It’s easy to be downloaded,” like, you know and she showed me how to do the app.’ *Participant 8*

> ‘They were very good actually, very good. Yes, very clear, very patient. I didn’t feel rushed to any of the tests, you know, all the stuff that we did that day in that first appointment, or indeed second one. You know, it didn’t feel rushed or anything. It was fine.’ *Participant 15*

However, some participants were concerned about forgetting the instructions and would have liked additional detailed instruction leaflets, for example how long to persevere to achieve an acceptable spirometer test result. Some participants suggested scheduled phone/face to face meetings from the clinical team. Examples include:

> ‘I was, say, 70, and I was blowing 110 times, if I didn’t know anything about it, I’d just think, oh my God, this is horrendous. So you end up doing more then, you see. And obviously the more you do, the worse you get, and it just has a knock-on effect then. But yes, just a bit of info, really, about the blowing I’d have thought, upfront before you take it home.’ *Participant 7*

> ‘It was the last bit. There was a lot to take in and trying to work it out and then you were left-Obviously, you’re going home and you’re doing it, on your own basically. So, if it was more simplified and it was written down on maybe an A5 saying, this is what you need to do, and you could read it when you’re at home, that would make it a lot - For my personal preference.’ *Participant 3*

*Building confidence and ongoing support:* participants described that the knowledge, approachability and support, particularly when seeking advice, of staff was important in developing their confidence and motivating to undertake home testing. Examples of support included:

> ‘It makes you feel more confident that you’re being more looked after. You know the aftercare is still there even though you’re at home. I think it’s important and it gives you the motivation to carry on doing the tests.’ Participant 3

> ‘I think it’s pretty self-explanatory, but you could call the team if you had questions, which I think was good. And they did check up on you once or twice. That’s when I told them my problems with the orange one, I didn’t get any accepted trials and they said that the app is sometimes really sensitive’. *Participant 14*

## Discussion

In line with the NHS Long Term Plan, ^26^ promoting community-based and patient-centred care, there is an increasing emphasis on shifting diagnostic services closer to home. Understanding patient perspectives on these evolving models of care is essential to ensure uptake and effectiveness. ^27, 28^ Our study explored patient experiences with home-based asthma diagnostic devices, including FeNO and spirometry. We found that home-based diagnostic approach is acceptable to patients in the asthma diagnostic setting, but perceived benefit and burden varied amongst individuals. Our findings highlighted the key motivators, barriers and enables of adopting digital health in the asthma diagnostic process. These findings form the basis for future service design and planning for using digital health products for asthma diagnosis. These insights are also transferable to other contexts, including the use of digital devices for home monitoring to support asthma self-management.

Consistent with previous studies, ^29–31^ we have demonstrated that home-based testing strategy using digital health devices empowers patient to be health proactive and actively engage in self-management of their asthma, providing sense of control of their diagnostic journey. Furthermore, the early involvement of patients in their diagnostic process may facilitate later shared decision-making, potentially leading to better clinician-patient partnership in disease management, improved patient satisfaction, thereby result in better asthma control, and overall improved quality of life. ^32^

We have demonstrated that a home-based digital healthcare system (including hand-held spirometer and FeNO devices) had the potential to revolutionise the self-management of asthma but adherence to twice-daily measurements fell within the first month of testing. ^20^ Longer duration and increased frequency of measurements represent an increased burden and forms a significant barrier to this testing strategy. Nevertheless, compared to long-term monitoring, home testing to establish diagnosis requires shorter term engagement (over 1-2 weeks) and likely to carry a better adherence rate.

Whilst many identified barriers are modifiable through careful design and planning (such as a user-friendly devices and app designs, ease of data capture, automated data sharing between healthcare professionals and patients, adequate training and support), others may be more challenging. For example, the feeling of lack of privacy and stigma of performing breathing tests in front of others and the challenges in breathing technique, particular for spirometry. Whilst the frequency and duration of home measurements may be minimised through future studies in determining the optimal timing of measurements, because spirometry is an effort dependent, forced expiratory technique, repeated blows are mandatory for quality control and cannot be avoided. These challenges may be addressed through AI-based virtual assistance and emerging models such as tele-coaching at home^33^.

We included a diverse group of patients with varying diagnostic outcomes, ages, occupations, and with a third from ethnic minority backgrounds. However, as with all qualitative studies the findings may not be representative of all patients with suspected asthma. Nonetheless, the diversity of baseline characteristics in our sample provides valuable insights into patient perspectives.

## Conclusion

The barriers and enablers to home-based asthma diagnosis using handheld spirometry and FeNO should be carefully considered and optimised during service implementation. The clinical and cost effectiveness should be evaluated as the next step.

## Supporting information

OLS

## Contributorship Statement

BK contributed to the data acquisition, analysis and writing of the submitted article. JS contributed to data analysis and writing of manuscript. RW and CSM contributed to the conception of the study, planning, set up, study oversight, writing and reviewing of the submitted article. SJF and AS contributed critical review to the study design and manuscript.

## Acknowledgement

We would like to thank the RADicA study participants for their time and commitment to the study and the RADicA study team for help with data collection. Binish Khatoon, Ran Wang, Clare Murray, Angela Simpson and Stephen Fowler are supported by the NIHR Manchester BRC. Ran Wang is supported by NIHR Clinical Lectureship.

## Data sharing statement

The authors will consider all reasonable requests for deidentified data, following approval by the study sponsors. Proposals should be directed to. To gain access, data requestors will need to sign a data access agreement.

