## Supplementary material for "What do patients think about home-based testing for better asthma diagnosis? - Insights from a qualitative study": OLS

**Dr Ran Wang**

2nd floor Education Research Centre

Wythenshawe Hospital

Southmoor road

Manchester

M23 9LT

### Detailed methods

Home spirometer (MIR Spirobank Smart, Intermedical, UK) and FeNO (NOBreath, Bedfont, UK) devices were provided. With permission, the app (MIR Spiro software) associated with the spirometers was downloaded, set up and installed on their smart phones in clinic. MIR Spiro software is compliant with the international ATS/ERS guidelines[24] and standards for spirometry. The software provides a virtual assistant for users to achieve optimum spirometry blow technique. In-person training and written instructions were provided for both devices.

Participants were asked to use FeNO and spirometer 4 times a day, e.g. 4-5 hourly (±1hr) when awake (e.g. 0600-0800, 1100-1300, 1600-1800, 2100-2400hr) and whenever symptomatic (before salbutamol use) for one week. For the second week, participants were asked to use spirometer twice-daily (0600-0800 and 2100-2400hr) and whenever symptomatic.

Whilst home spirometry results and flow volume loop were obtained through direct emailing of app-generated PDF reports to the study team, participants were asked to write down FeNO results on a diary book, along with symptoms experienced during home monitoring period. Participants were also asked to record in the diary when tests were completed and reasons for missed tests. The diary book also contained simple written instructions of how to operate the devices, helpline and short explanations of what FeNO and spirometry measures and simple guide for how to interpret results. Reminders were sent via text messages or emails, with participant permission, on day 3 and 5; with a follow up phone call on day 7. Remote telephone support was available throughout the monitoring period. Participants were asked to return the devices and diaries following the home monitoring period.

Participants were invited to take part in a semi-structured interview at the time of recruitment (prior to the 2-week home monitoring period); interviews were undertaken in-person or via telephone, according to participants preference, a maximum of 3 weeks following the monitoring period. An interview topic guide was developed with the input from our public and participant involvement and engagement activities and the wider study team (**Table E1**). Interviews were conducted in-person or *via* telephone, according to participants preference, and undertaken by BK, an experienced qualitative methodologist who was independent from the clinical research team. The interviewer had no specific prior knowledge or background in asthma or related health topics. The researcher was not involved in recruiting patients to the clinical study or in training patients to use the home diagnostic devices. With participants permission the interviews were recorded, transcribed anonymously and analysed. The semi-structured interview style was adopted to ensure the aims of the study were addressed and focused on patients' recent experiences of using the domiciliary diagnostic devices, the perceived benefits and challenges of home testing. Prompts were used to expand and clarify, if needed. Field notes were used to document initial reactions, emotions and cues which were not possible to capture on the recorder. The interviewer discussed the decision to stop further interviews with the wider team when it was felt that further interviews would not yield new insights or themes reaching data saturation; data collection was stopped when data saturation was reached and wider team agreed.

**Table E1: Interview topic guide**

| **Topic** | **Prompts/questions** |
|---|---|
| Introductions and background | |
| *Introductions*<br><br>*Opening question*<br><br>How have your 'asthma' symptoms been? | Welcome, check purpose of this interview<br><br>Obtain consent and to audio recording the interview |
| Experience of using home diagnostic devices | |
| Thinking back to using asthma testing devices, can you describe how you found using them?<br><br>Were they easy to use? If so why? If not why not? | Ask about each devise separately:<br>• Spirometry<br>• FeNO<br><br>Ask about the design of the devices, ease of use, |

| | |
|---|---|
| Did you manage to complete all the test?<br><br>What were your opinions of the written and verbal Instructions/training offered? | and any difficulties experienced<br>Depending on answer: Can you explain what motivated you to ensure you completed all test? or Can you outline the reasons you were unable to complete all measures?<br>Did you access to support |
| Perceived benefit or what's good about home diagnosis | |
| What do you think about using the home diagnostic devices?<br><br>From your point of view how important would is home testing be if it was part of your usual care? | What is good about the home diagnosis?<br>Do you feel it is an acceptable way to assess if people they have asthma? Can you explain your answers.<br><br>What would encourage you to undertake home testing? |
| Perceived drawbacks or what's not good about home diagnosis | |
| Do you think there are any drawbacks to home diagnosis testing? | If home testing was part of your usual care what would prevent you to undertake the testing? |
| Future improvement | |
| If you suggest 3 things we could improve on, what would they be? | In an ideal situation what do you think would help to ensure effective home testing? |
| **Closing the interview** | Thank participant for their time, explain what happen to the results and ascertain whether they would like feedback |
